# Clinical Outcomes With the Use of Prophylactic Versus Therapeutic Anticoagulation in COVID-19

**DOI:** 10.1101/2020.07.20.20147769

**Authors:** Jishu K. Motta, Rahila O. Ogunnaike, Rutvik Shah, Stephanie Stroever, Harold V. Cedeño, Shyam K. Thapa, John J. Chronakos, Eric J. Jimenez, Joann Petrini, Abhijith Hegde

## Abstract

**Background:** This study is the first of its kind to assess the impact of preemptive therapeutic dose anticoagulation on mortality compared to prophylactic anticoagulation among COVID-19 patients. Its findings provide insight to clinicians regarding the management of COVID-19, particularly with the known prothrombotic state.

**Research Question:** To determine the impact of anticoagulation on in-hospital mortality among COVID-19 positive patients with the *a priori* hypothesis that there would be a lower risk of in-hospital mortality with use of preemptive therapeutic over prophylactic dose enoxaparin or heparin.

**Study Design and Methods:** *Study Design:* Retrospective cohort study from April 1 - April 25, 2020. The date of final follow-up was June 12, 2020.

*Setting:* Two large, acute care hospitals in Western Connecticut.

*Participant:* Five hundred and one inpatients were identified after discharge as 18 years or older and positive for SARS-CoV-2. The final sample size included 374 patients after applying exclusion criteria. Demographic variables were collected via hospital billing inquiries, while the clinical variables were abstracted from patients’ medical records.

*Exposure:* Preemptive enoxaparin or heparin at a therapeutic or prophylactic dose.

*Main Outcom:* In-hospital mortality.

*Results:* When comparing preemptive therapeutic to prophylactic anticoagulation through multi-variable analysis, risk of in-hospital mortality was 2.3 times greater in patients receiving preemptive therapeutic anticoagulation (95% CI = 1.0, 4.9; p = 0.04).

*Interpretation:* An increase in in-hospital mortality was observed with preemptive therapeutic anticoagulation. Thus, in the management of COVID-19 and its complications, we recommend further research and cautious use of preemptive therapeutic over prophylactic anticoagulation.

## Introduction

The first outbreak of SARS-CoV-1 occurred 18 years ago. Scientists identified bats as the primary reservoir and additional SARS-related coronaviruses have been discovered among humans since that time.^1,2^ In December 2019, a new beta-coronavirus, SARS-CoV-2, infected the first humans in Wuhan, China and rapidly spread throughout the world creating a global pandemic.^3^ The disease, also known as COVID-19, has caused extensive infection and mortality worldwide with over 2 million cases and over 115,000 deaths in the United States as of June 18, 2020.^4^ The trigger for severe disease and the extent of physiological damage in the human body is currently unknown, though there are some similarities with other viruses in the *Coronaviridae* family.^1,2,5^ SARS-CoV-2 uses the angiotensin-converting enzyme 2 (ACE2) for cell entry into type II pneumocytes which results in diffuse alveolar damage and acute lung injury.^6–10^ Patients with severe illness have also been noted to have elevated d-dimer, C-reactive protein (CRP), and interleukin-6 (IL-6) levels.^11–18^ Additionally, there is evidence that patients develop thrombotic complications such as pulmonary embolism and unexplained renal failure.^11–22^ As observed with SARS-CoV-1, infections that elicit a systemic inflammatory response can cause an imbalance between procoagulant and anticoagulant homeostatic mechanisms.^11^

Postmortem analyses of patients with SARS viruses revealed microthrombi in pulmonary microvasculature, deep vein thrombosis, and pulmonary embolisms.^11,22^ There have also been reports of COVID-19-induced chilblains and acute acro-ischemia in younger patients.^21^ Prior studies sought to determine possible benefits in treating COVID-19 patients with anticoagulation (AC) therapy. Shi et al. found that 42 patients had a significant decrease in the proportion of lymphocytes, d-dimer levels, and fibrinogen degradation products among a small sample of patients that received prophylactic low molecular weight heparin (LMWH), with no decrease in length of hospitalization.^6^ Tang et al. found no difference in 28-day mortality among a sample of 449 COVID-19 patients who did or did not receive prophylactic LMWH. However, a sub-group analysis among patients with markers for severe disease revealed a significantly lower rate of mortality among patients who received heparin.^23^ Paranjpe and Fuster et al described an association between the use of therapeutic AC and decreased length of stay (LOS) but no significant change in in-hospital mortality compared to patients who did not receive AC. However, in patients that were mechanically ventilated and treated with therapeutic AC, they described a significant decrease in mortality and LOS compared to no AC.^24^

All of these findings support the hypothesis that COVID-19 can induce a prothrombotic state. Despite some evidence, additional research is needed to identify possible treatments to minimize the potential harm among patients who are at risk for thrombotic complications. The primary objective of this study was to determine the impact of AC on the mortality among patients who received a therapeutic versus prophylactic dose of enoxaparin or heparin. Based on prior studies that showed a prothrombotic state induced by COVID-19 leading to increased mortality, we hypothesize that preemptive treatment with therapeutic AC can lower this risk.

The secondary objective was to determine the difference between the two groups in in-hospital mortality, among patients with a peak CRP ≥ 200 mg/L.

This study is the first of its kind to compare clinical outcomes of COVID-19 positive patients who received preemptive therapeutic versus prophylactic AC, started upon admission as opposed to for the treatment of a thrombotic condition. Previous studies assessed the impact of any AC therapy compared to none. This study can provide guidance to clinicians on the appropriate utilization of AC in COVID-19 patients.

## Methods

### Study Design, Setting, and Participants

We conducted a retrospective cohort study of patients from two large, acute care hospitals in Western Connecticut. Both hospitals are located in Fairfield County, which had the highest incidence of infection in Connecticut with more than 24,000 cases of the virus to date^25^. The protocol and subsequent work were granted exempt status by the Biomedical Research Alliance of New York (BRANY) Institutional Review Board under category #4(iii), as detailed in 45 CFR 46.104(d) and BRANY’s Standard Operating Procedure.

The study included adult patients admitted with a diagnosis of COVID 19 (ICD-10 code B97.29, J12.89, J18.9, U07.1) between April 1 and April 25, 2020, and treated with AC during their inpatient stay. AC was defined as the therapeutic or prophylactic use of enoxaparin or heparin, both regimens started preemptively upon admission. Patients were excluded if they did not take enoxaparin or heparin during their inpatient stay or if they were on other forms of AC prior to or during their hospitalization. Demographic variables were collected via hospital billing inquiries, while the clinical variables were abstracted from patients’ medical records.

## Variables

### Main Outcomes and Predictors

The primary outcome measures were a dichotomous variable for death. The secondary outcome measure was mortality in patients with peak ≥ 200 mg/L.

The primary exposure variable was dose of AC. Therapeutic dosage for enoxaparin was defined as 1 mg/kg subcutaneously twice daily or 1.5 mg/kg subcutaneously daily or based on renal function, or higher doses titrated to anti-Factor Xa range of 0.6 to 1 IU/mL (for twice daily dosing) and 1 to 2 IU/mL (for daily dosing).^26^ Prophylactic dosage for enoxaparin was defined as 30 or 40 mg subcutaneously every day. For heparin, therapeutic dosage was defined as intravenous heparin titrated to an activated partial thromboplastin time (aPTT) between 70 and 110 sec, and prophylactic dosage was defined as 5000 units given subcutaneously every 8 hours. Patients were assigned to the therapeutic group if they preemptively received a therapeutic dosage of either medication at any time or the prophylaxis group if they only received prophylaxis for the duration of their inpatient stay. Patients who received therapeutic anticoagulation specifically for a thrombotic indication were excluded from this study. We recognize that the dichotomization of a time-varying variable may introduce some bias to the analysis. Thus, we created an alternative exposure variable for sensitivity analyses that defined AC as prophylactic or therapeutic dosage at time of admission.

### Covariates

Numerous patient and treatment-related variables were included as possible confounders given their potential association with the outcomes (Table 1). Demographic variables included age, gender, race, and ethnicity. Race was defined as White, Black or African American, and others. Ethnicity was defined as either Hispanic or non-Hispanic. We also included body mass index, smoking status (never/ever), diabetes, current immunosuppression, prior history of heart disease, pulmonary disease, kidney disease, cancer, and hyperlipidemia. History of heart disease was defined as a dichotomous variable for any of the following: hypertension, congestive heart failure, myocardial infarction, atrial fibrillation, and other heart disease. History of any pulmonary disease included asthma, chronic obstructive pulmonary disease, pulmonary embolism, obstructive sleep apnea, and other.

**Table 1.**
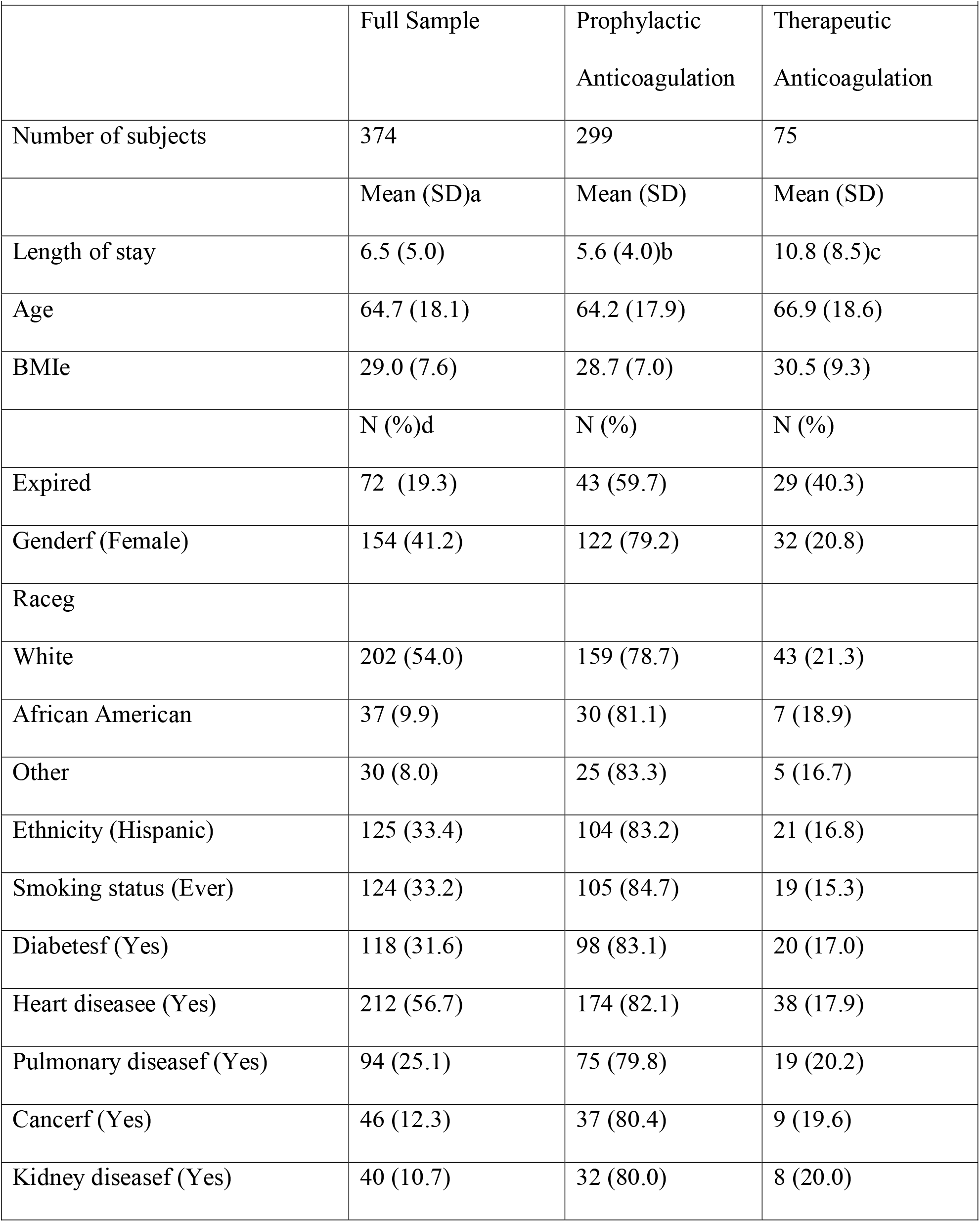
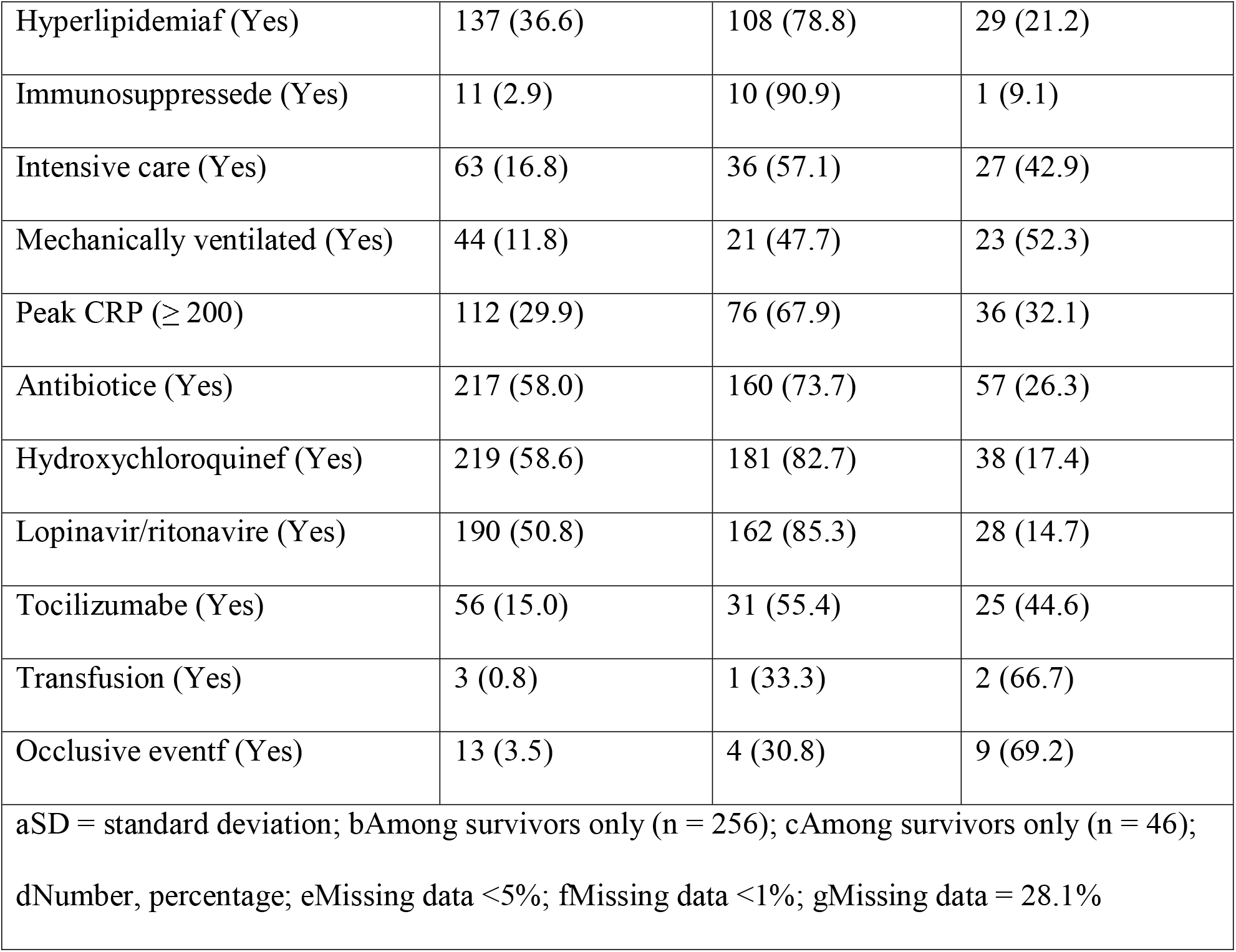
Descriptive statistics among COVID-19 positive patients from two hospitals in Western Connecticut in the COV-AC study.

History of cancer and kidney disease were each a dichotomous variable reflecting either of these conditions. We included immunosuppression as a dichotomous variable that reflected diseases such as transplant, myelodysplasia, rheumatoid arthritis, psoriatic arthritis, and cancer. Cancer and immunosuppression had some overlap in patients. However, they were maintained as separate variables, because the correlation between the variables was less than 0.25.

Treatment-related variables included the need for ICU admission, mechanical ventilation, and treatment with antibiotics, Tocilizumab, Hydroxychloroquine, or Lopinavir/Ritonavir. We also included a dichotomous variable for peak CRP defined as < 200 mg/L or ≥ 200 mg/L to reflect severity. Additionally, we collected data on the outcome of receiving pRBC transfusion and the occurrence of an arteriovenous occlusive event.

### Statistical methods

We performed all analyses with StateSE 16. We computed descriptive statistics as percentages of the total for dichotomous and categorical variables and mean with standard deviation for continuous. We also used Independent Student’s t-test to assess group differences for each continuous covariate and chi-square or Fisher’s exact test for dichotomous/categorical covariates. We accounted for missing data with list wise deletion, and set the alpha for statistical hypothesis testing at 0.05.

We used a multi-variable logistic regression model to determine risk differences in mortality given AC dosage. We developed the model according to *a priori* hypotheses on confounders that were both associated with our primary exposure and outcome and not in the causal pathway. We also verified model assumptions. We performed a sensitivity analysis and qualitatively assessed differences in effect size and direction with the alternative exposure variable. We used similar methods to assess differences in mortality among patients with CRP ≥200 mg/L.

We performed a *post hoc* investigation of the cause of death among patients in our cohort to explore the reason for the increased risk of death among patients on therapeutic dosage of anticoagulant. We computed descriptive statistics on the primary cause of death and election of comfort measures among all patients that expired.

Lastly, we assessed the difference in receiving pRBC transfusion and incidence of occlusive events among all patients on therapeutic and prophylactic AC with univariate logistic regression.

## Results

### Participants

A total of 501 inpatients were initially identified as 18 years or older and positive for SARS-CoV-2. A total of 374 patients were included in this study following the application of exclusion criteria (Figure 1). There was limited missing data with most variables missing less than 1 to 5 percent (Table 1). Race had more than 20 percent missing data, because Hispanic was listed as race in some medical records. Thus, the race of the patient was unknown. List wise deletion for the multi-variable logistic model resulted in a final sample size of 351. The sub-group analysis to determine risk of mortality among patients with CRP ≥ 200 mg/L included 104 patients.

We provide descriptive statistics in Table 1. The average age was 64.7 years old, more than half of the sample was male (58.6%), and the majority was White (54.0%) and non-Hispanic (63.6%). Nearly all patients in our sample took enoxaparin at some time during their inpatient stay (93.5%), while less than one-fifth took heparin (14.8%). Some patients took both medications at different times, depending on their treatment requirements. Seventy-five (20.1%) patients were on therapeutic AC, and seventy-two patients expired (19.2%).

### Risk differences in mortality

There was a statistically significant increase in the risk of mortality in the therapeutic AC group compared with the prophylactic AC group upon crude analysis (Table 2). The full logistic model included AC dosage, age, ethnicity, diabetes, history of cancer or heart disease, hyperlipidemia, peak CRP, intensive care, mechanical ventilation, and antibiotic use.

**Table 2.**
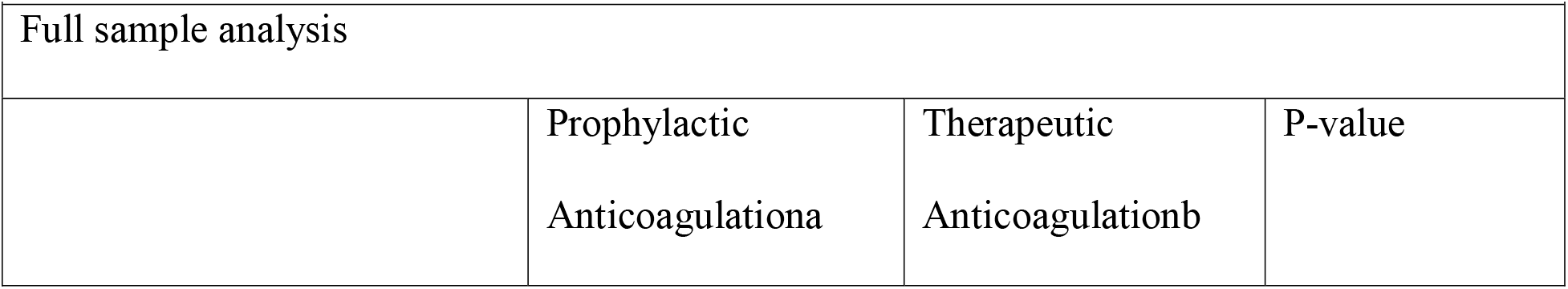
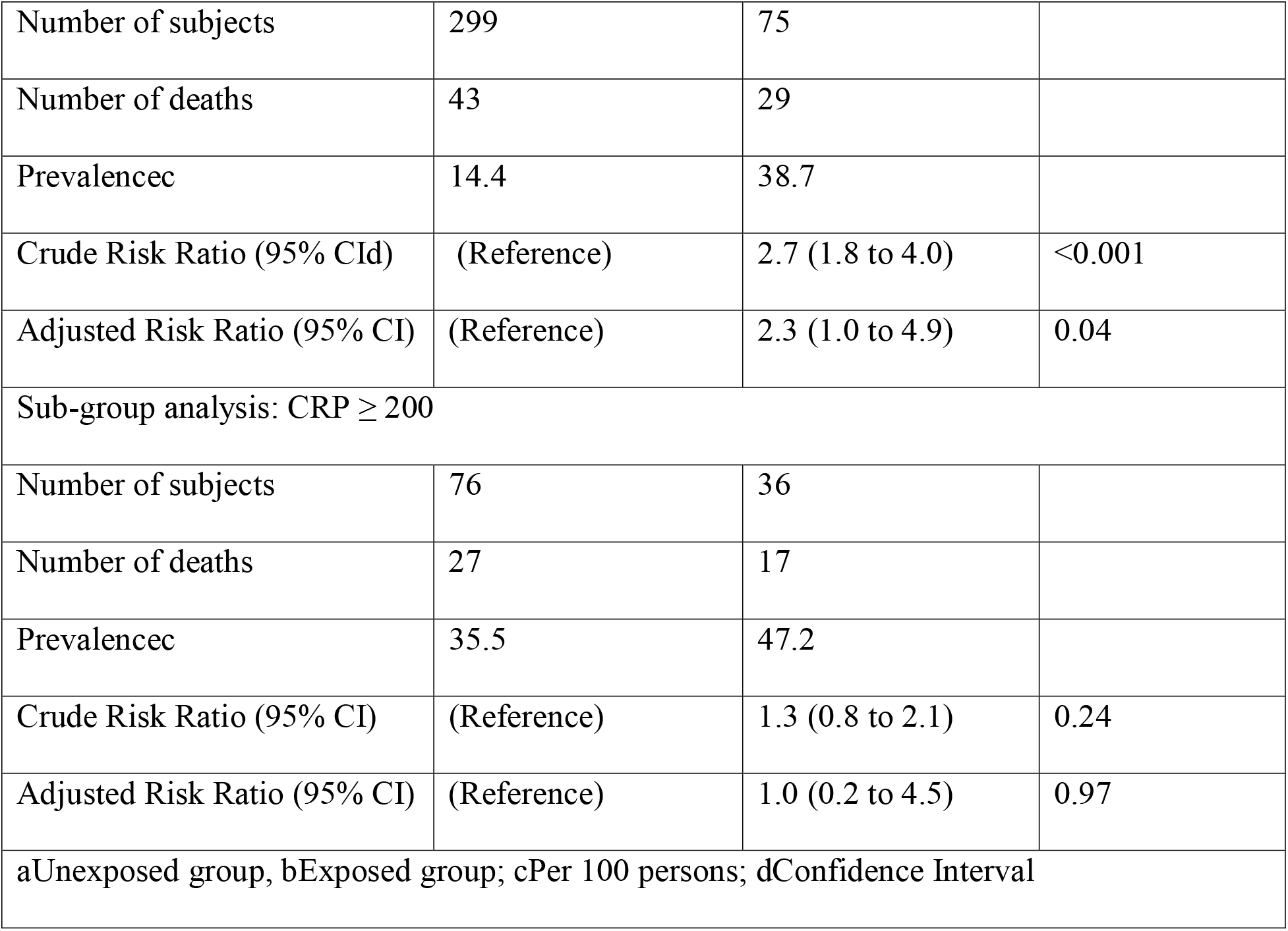
Results from multi-variable logistic regression to determine the difference in risk of mortality between patients on therapeutic versus prophylactic doses of anticoagulation in the COV-AC full sample and among patients with peak C-reactive protein ≥ 200 (α = 0.05)

The risk of mortality was higher (aRR = 2.3, 95% CI = 1.0, 4.9, p = 0.04) for patients on therapeutic AC when compared to prophylactic AC after controlling for all variables in the model (Table 2).

### Sub-group analyses

We performed a sub-group analysis of patients with a CRP ≥ 200 mg/L to determine if there was a risk difference in mortality. We hypothesized that patients with evidence of severe disease may benefit from therapeutic AC. However, multi-variable logistic regression that included AC dosage, age, ethnicity, diabetes, history of cancer, history of any heart disease, intensive care, mechanical ventilation, and use of antibiotics, hydroxychloroquine, and tocilizumab demonstrated no difference in the risk of mortality between patients on therapeutic and prophylactic AC (aRR = 1.0, 95% CI = 0.2, 4.5, p = 0.97) (Table 2).

Additionally, there was no difference found in the transfusion of pRBC or incidence of arteriovenous occlusive events across AC dosage on univariate analysis.

### Sensitivity analyses

Qualitatively, we found there was little difference in the risk ratios in the sensitivity analyses for risk of mortality. However, the confidence intervals in the primary exposure variable were more precise than the alternative exposure variable. The conclusions remain the same regardless of the definition for therapeutic AC.

### *Post hoc* analyses

The main causes of death among patients that expired are provided in Table 3. The majority of patients expired due to worsening oxygenation (71.8%) and acute respiratory failure with hypoxia. Patients also expired due to shock and multi-organ failure. Approximately 8% of patients died from other causes, including anoxic brain injury due to hemorrhage (n = 1), kidney dysfunction with inability to access hemodialysis port (n = 1), and failure to thrive with encephalopathy (n = 1). Additionally, 64.4% of patients elected to receive comfort measures only for end of life care.

**Table 3.** Causes of death among COVID-19 positive patients from two hospitals in Western Connecticut in the COV-AC studya

|  |  |  |  |
| --- | --- | --- | --- |
|  | Expired | Prophylactic Anticoagulation | Therapeutic Anticoagulation |
| Number of subjects | 72 | 43 | 29 |
|  | N (%)<sup>b</sup> | N (%) | N (%) |
| Acute respiratory failure with hypoxia | 51 (70.8) | 31 (72.1) | 20 (68.9) |
| Multi-organ failure and septic shock | 5 (6.9) | 3 (7.0) | 2 (6.9) |
| Cardiac arrest | 9 (12.5) | 6 (13.9) | 3 (10.3) |
| Anoxic brain injury due to hemorrhage | 1 (1.4) | 0 (0.0) | 1 (100.0) |
| Kidney injury and failure to access hemodialysis port | 1 (1.4) | 1 (100.0) | 0 (0.0) |
| Failure to thrive with encephalopathy | 1 (1.4) | 0 (0.0) | 1 (50.0) |
| aCauses of death are not mutually exclusive. Some patients died of multiple indications;<br>bNumber, percentage; |  |  |  |

## Discussion

Since the onset of the COVID 19 pandemic, aggressive efforts to elucidate an effective anti-viral or biologic agent against this virus, have yielded inconclusive data^27^. Giannis et al and Mehta et al have discussed the hyper inflammatory state that occurs in patients with COVID-19.^9,11^ Postmortem reports of thrombo-embolic disease in COVID-19 patients indicate systemic thromboses possibly leading to the clinical decline seen in these patients.^11,22^ Our study evaluated whether preemptive administration of prophylactic AC versus therapeutic AC resulted in a difference in in-hospital mortality, among all COVID 19 patients and among a subset with significantly elevated CRP levels.

Among all in-patients with COVID-19, after controlling for relevant covariates, in-hospital mortality was 2.3 times higher in preemptive therapeutic over prophylactic anticoagulation. When we focused on patients with greater severity of disease (CRP ≥200), there was still no clinical improvement in outcomes of preemptive therapeutic anticoagulation. Although these results were different from the study by Paranjpe and Fuster et al, this study compared prophylactic with therapeutic AC, while adjusting for multiple confounders. Klok et al also did not find a difference in mortality in COVID-19 patients treated with therapeutic anticoagulation at baseline, despite reduction in thrombotic events^28^. The presence of a COVID-19 induced immunothrombotic state has been suggested in the literature, and disseminated intravascular coagulation, which can account for thrombosis on a consumptive basis, has been reported in autopsy specimens^29^. Since the thrombotic effects of COVID 19 are not completely understood, it may very well be the case that therapeutic anticoagulation is an ineffective treatment with worse clinical outcomes for this syndrome. When cause of death was evaluated, it was found that most patients expired due to refractory acute respiratory failure with hypoxia as well as shock and multi-organ system failure. Although thrombosis could have played a role in these top causes of death, they can also be attributed to other reasons unrelated to thrombosis, such as direct end organ damage due to the virus or to the systemic inflammatory response syndrome caused by the virus. Regardless it does not seem from our analyses that therapeutic dosing of anticoagulation prevented overall disease progression. Of note the high incidence of comfort measures in this study most likely reflects a transition after the disease had worsened to the point where further care would not have been beneficial to the patient, and given the high risk of cardiopulmonary resuscitation to healthcare workers and the poor outcomes in COVID, many times patients were transitioned after other aggressive interventions were failing and death was imminent in an effort to avoid it.

There were several limitations recognized in this study. First, this is a retrospective study that did not randomize patients to treatment groups. Group assignments may be biased by prescriber preferences or differences in unmeasured clinical variables that may bias the effect sizes of these variables. Second, clinical outcomes, such as mortality after the patients left the hospital, were not captured in this study. Therefore, there may be some misclassification bias in the outcome. Additionally, we included variables in our model that we hypothesized would impact mortality among patients on AC. However, it is likely there is unmeasured confounding present in the model.

The patients in this sample were selected from two institutions in Western Connecticut and were predominantly older, non-Hispanic, and White. Interpretation of the results should only be generalized to similar patient populations. Future research should utilize a randomized control design and include additional factors related to mortality, bleeding, and complications of AC utilization.

## Conclusion

Among hospitalized patients with COVID-19, an increase in the risk of mortality was observed in patients who preemptively received therapeutic AC when compared to those who received prophylactic AC. It is therefore important to consider the risks and benefits for the patient as well as the healthcare system when using preemptive therapeutic AC in COVID-19. We recommend further research and cautious use of therapeutic AC over standard prophylactic AC in the management of COVID-19 and its complications.

## Data Availability

Data analyzed in this manuscript is available upon request from the corresponding author, Jishu Kaul Motta, at

## Acknowledgments

Abhijith Hegde, MD and Jishu K. Motta, MD take responsibility for the integrity of the work as a whole, from inception to published article.

Rutvik Shah, MD, Jishu K. Motta, MD and Rahila O. Ogunnaike, MD, were involved in conception and design of the research study, data acquisition, drafting and revision of the manuscript. Harold V. Cedeno, MD and Shyam K. Thapa, MD were involved in data acquisition, drafting and revision of the manuscript. Stephanie Stroever, PhD was involved in data analysis and interpretation, writing of the methods and results sections, and revision of the manuscript. Joann Petrini, PhD, MPH, John J. Chronakos, MD, Eric J. Jimenez, MD and Abhijith Hegde, MD were involved in critical revision of the manuscript for important intellectual content, and final approval of the version to be published.

No external funding was received for this study.

The authors wish to acknowledge the following contributors to this work, who did not receive any specific compensation for their contributions. From the Department of Pulmonary and Critical Care, Danbury Hospital: Dr. Thomas Botta, Dr. Guillermo Ballarino, Dr. Jose Mendez, Dr. Loren Inigo-Santiago and Dr. Dmitriy Golovyan. From the Department of Infectious Diseases, Danbury Hospital: Dr. Paul Nee, Dr. John Stratidis, Dr. Gary Schleiter and Dr. Martha DesBiens. From the Department of Research and Innovation, Nuvance Health: Jon Serino.

## Notes

### Competing Interest Statement

The authors have declared no competing interest.

### Author Declarations

The protocol and subsequent work were granted exempt status by the Biomedical Research Alliance of New York (BRANY) Institutional Review Board under category #4(iii), as detailed in 45 CFR 46.104(d) and BRANYs Standard Operating Procedure.

## References

1. Zhou P, Yang X-L, Wang X-G, et al. A pneumonia outbreak associated with a new coronavirus of probable bat origin. Nature. 2020;579(7798):270–273. doi:10.1038/s41586-020-2012-7

2. Zhu N, Zhang D, Wang W, et al. A Novel Coronavirus from Patients with Pneumonia in China, 2019. New England Journal of Medicine. 2020;382(8):727–733. doi:10.1056/NEJMoa2001017

3. WHO Director-General’s remarks at the media briefing on 2019-nCoV on 11 February 2020. Accessed April 11, 2020. https://www.who.int/dg/speeches/detail/who-director-general-s-remarks-at-the-media-briefing-on-2019-ncov-on-11-february-2020.

4. WHO Coronavirus Disease (COVID-19) Dashboard. Accessed May 10, 2020. https://covid19.who.int/.

5. Thakkar N, Yadavalli T, Jaishankar D, Shukla D. Emerging Roles of Heparanase in Viral Pathogenesis. Pathogens. 2017;6(3):43. doi:10.3390/pathogens6030043

6. Shi C, Wang C, Wang H, et al. The Potential of Low Molecular Weight Heparin to Mitigate Cytokine Storm in Severe COVID-19 Patients: A Retrospective Clinical Study. Pharmacology and Therapeutics; 2020. doi:10.1101/2020.03.28.20046144

7. Gralinski LE, Bankhead A, Jeng S, et al. Mechanisms of Severe Acute Respiratory Syndrome Coronavirus-Induced Acute Lung Injury. Dermody TS, ed. mBio. 2013;4(4). doi:10.1128/mBio.00271-13

8. McGonagle D, O’Donnell JS, Sharif K, Emery P, Bridgewood C. Immune mechanisms of pulmonary intravascular coagulopathy in COVID-19 pneumonia. The Lancet Rheumatology. Published online May 2020:S2665991320301211. doi:10.1016/S2665-9913(20)30121-1

9. Mehta P, McAuley DF, Brown M, Sanchez E, Tattersall RS, Manson JJ. COVID-19: consider cytokine storm syndromes and immunosuppression. The Lancet. 2020;395(10229):1033–1034. doi:10.1016/S0140-6736(20)30628-0

10. Yuki K, Fujiogi M, Koutsogiannaki S. COVID-19 pathophysiology: A review. Clin Immunol. 2020;215:108427. doi:10.1016/j.clim.2020.108427

11. Giannis D, Ziogas IA, Gianni P. Coagulation disorders in coronavirus infected patients: COVID-19, SARS-CoV-1, MERS-CoV and lessons from the past. Journal of Clinical Virology. 2020;127:104362. doi:10.1016/j.jcv.2020.104362

12. Goyal P, Choi JJ, Pinheiro LC, et al. Clinical Characteristics of Covid-19 in New York City. N Engl J Med. Published online April 17, 2020:NEJMc2010419. doi:10.1056/NEJMc2010419

13. Huang C, Wang Y, Li X, et al. Clinical features of patients infected with 2019 novel coronavirus in Wuhan, China. The Lancet. 2020;395(10223):497–506. doi:10.1016/S0140-6736(20)30183-5

14. Richardson S, Hirsch JS, Narasimhan M, et al. Presenting Characteristics, Comorbidities, and Outcomes Among 5700 Patients Hospitalized With COVID-19 in the New York City Area. JAMA. Published online April 22, 2020. doi:10.1001/jama.2020.6775

15. Tang N, Li D, Wang X, Sun Z. Abnormal coagulation parameters are associated with poor prognosis in patients with novel coronavirus pneumonia. Journal of Thrombosis and Haemostasis. 2020;18(4):844–847. doi:10.1111/jth.14768

16. Terpos E, Ntanasis-Stathopoulos I, Elalamy I, et al. Hematological findings and complications of COVID-19. American Journal of Hematology. Published online April 13, 2020. doi:10.1002/ajh.25829

17. Wu Z, McGoogan JM. Characteristics of and Important Lessons From the Coronavirus Disease 2019 (COVID-19) Outbreak in China: Summary of a Report of 72 314 Cases From the Chinese Center for Disease Control and Prevention. JAMA. 2020;323(13):1239–1242. doi:10.1001/jama.2020.2648

18. Yan Z, Wei C, Meng X, et al. Clinical and coagulation characteristics of 7 patients with critical COVID-2019 pneumonia and acro-ischemia. Chinese Journal of Hematology. 2020;41(00):E006–E006. doi:10.3760/cma.j.issn.0253-2727.2020.0006

19. Danzi GB, Loffi M, Galeazzi G, Gherbesi E. Acute pulmonary embolism and COVID-19 pneumonia: a random association? Eur Heart J. doi:10.1093/eurheartj/ehaa254

20. Porfidia A, Pola R. Venous thromboembolism in COVID-19 patients. Journal of Thrombosis and Haemostasis. Published online April 15, 2020. doi:10.1111/jth.14842

21. Suarez-Valle A, Fernandez-Nieto D, Diaz-Guimaraens B, Dominguez-Santas M, Carretero I, Perez-Garcia B. Acro-ischemia in hospitalized COVID-19 patients. J Eur Acad Dermatol Venereol. Published online May 7, 2020. doi:10.1111/jdv.16592

22. Wichmann D, Sperhake J-P, Lütgehetmann M, et al. Autopsy Findings and Venous Thromboembolism in Patients With COVID-19: A Prospective Cohort Study. Ann Intern Med. Published online May 6, 2020. doi:10.7326/M20-2003

23. Tang N, Bai H, Chen X, Gong J, Li D, Sun Z. Anticoagulant treatment is associated with decreased mortality in severe coronavirus disease 2019 patients with coagulopathy. Journal of Thrombosis and Haemostasis. 2020;18(5):1094–1099. doi:10.1111/jth.14817

24. Paranjpe I, Fuster V, Lala A, et al. Association of Treatment Dose Anticoagulation with In-Hospital Survival Among Hospitalized Patients with COVID-19. J Am Coll Cardiol. Published online May 6, 2020. doi:10.1016/j.jacc.2020.05.001

25. Coronavirus. Accessed May 20, 2020. https://portal.ct.gov/Coronavirus

26. Wei MY, Ward SM. The anti-factor Xa range for low molecular weight heparin thromboprophylaxis. Hematology Reports. 2015;7(4). doi:10.4081/hr.2015.5844

27. Rameshrad M, Ghafoori M, Mohammadpour AH, Nayeri MJD, Hosseinzadeh H. A comprehensive review on drug repositioning against coronavirus disease 2019 (COVID19). Naunyn-Schmiedeberg’s Arch Pharmacol. Published online May 19, 2020. doi:10.1007/s00210-020-01901-6

28. F.A. Klok, M.J.H.A. Kruip, N.J.M. van der Meer, M.S. Arbous, D.A.M.P.J. Gommers, K.M. Kant, F.H.J. Kaptein, J. van Paassen, M.A.M. Stals, M.V. Huisman, H. Endeman. Incidence of thrombotic complications in critically ill ICU patients with COVID-19 Thromb. Res. (2020), doi:https://doi.org/10.1016/j.thromres.2020.04.041

29. S.E. Fox, A. Akmatbekoy, J.L. Harbert, et al. Pulmonary and Cardiac Pathology in Covid-19: The First Autopsy Series From New Orleans MedRxiv preprint (2020). doi:10.1101/2020.04.06.20050575

30. Li R, Rivers C, Tan Q, Murray MB, Toner E, Lipsitch M. Estimated Demand for US Hospital Inpatient and Intensive Care Unit Beds for Patients With COVID-19 Based on Comparisons With Wuhan and Guangzhou, China. JAMA Netw Open. 2020;3(5):e208297–e208297. doi:10.1001/jamanetworkopen.2020.8297

